# Effect of using Diffusion-Weightage Imaging on Diagnostic Accuracy of MRI in catching Parotid carcinoma: A meta-analysis

**DOI:** 10.1101/2023.06.27.23291962

**Authors:** Dev Desai, Marko Todorovski, Maria Eleni Malafi, Abhijay B. Shah, Dev Andharia

## Abstract

**Background:** A wide variety of benign and malignant tumors can develop in the salivary gland, which makes up roughly 3-5% of all head and neck tumors. Making a precise pre-operative distinction between benign and malignant salivary gland tumors, or identifying the precise histologic subtype, is crucial since this knowledge has a significant impact on the treatment strategy. The results of fine-needle aspiration cytology can occasionally be inaccurate since it is an intrusive procedure for diagnosing parotid tumors before surgery. Because of the tiny sample size or the deep placement of the tumor, it is sometimes impossible to gather adequate samples therefore Salivary gland tumors are increasingly being evaluated before surgery using MRI since it can pinpoint their precise location, size, and surrounding tissues.

**Methods:** Medical literature was comprehensively searched and reviewed without restrictions to particular study designs, or publication dates using PubMed, Cochrane Library, and Google Scholar databases for all relevant literature. The extraction of necessary data proceeded after specific inclusion and exclusion criteria were applied. The inclusion criteria were as follows:; (1) Literature that provided information about the accurate diagnosis with MRI(DWI) and only MRI ; (2) papers published in English; (3) papers with appropriate patient criteria. The exclusion criteria were: (1) articles that were not full text, (2) unpublished articles, and (3) non-English articles. In this Meta-Analysis, a total of 8 RCTs with a total of 944 patients were selected, out of which 3 RCTs with a total of 356 patients were selected for MRI and 5 RCTs with a total of 588 patients were selected for MRI+DWI study. wherein two writers independently assessed the caliber of each study as well as the use of the Cochrane tool for bias risk apprehension. The statistical software packages RevMan (Review Manager, version 5.3), SPSS (Statistical Package for the Social Sciences, version 20), and Excel in Stata 14 were used to perform the statistical analyses.

**Results:** We calculated the sensitivity and specificity of MRI in diagnosing parotid gland lesions in the different papers, For the MRI, The sensitivity is 0.81 with a CI of 95% in a range of 0.77 to 0.86, the mean being 0.049. The Sensitivity of the MRI is 0.790 with a CI of 95% in a range of (0.772 to 0.809) the mean being (0.0183). The Specificity of the MRI is 0.794 with a CI of 95% in a range of (0.613 to 0.975) the mean being (0.181). For the MRI+DWI study, The Sensitivity of the MRI+DWI is 0.618 with a CI of 95% in a range of (0.498 to 0.737); the mean being (0.119). The Specificity of the MRI+DWI is 0.892 with a CI of 95% in a range of (0.813 to 0.97) the mean being (0.079).

**Conclusion:** In addition to simple MRI or Single DWI MRI, A combination of both would detect higher lesions and it should be helpful for staging and early diagnosis purposes only.

## Introduction

A wide variety of benign and malignant tumors can develop in the salivary gland, which makes up roughly 3-5% of all head and neck tumors. [1] Making a precise pre-operative distinction between benign and malignant salivary gland tumors, or identifying the precise histologic subtype, is crucial since this knowledge has a significant impact on the treatment strategy. [2] Malignant tumors are typically treated with a more aggressive approach, such as a total parotidectomy that may involve removing the facial nerve, whereas benign tumors are advised to undergo local excision or superficial parotidectomy. [3] [4]

For proper surgical intervention, timely diagnosis plays a crucial role. Fine needle aspiration cytology is one of the many diagnostic modalities that can be used. However, it has a few disadvantages.

The results of fine-needle aspiration cytology can occasionally be inaccurate since it is an intrusive procedure for diagnosing parotid tumors prior to surgery. Because of the tiny sample size or the deep placement of the tumor, it is sometimes impossible to gather adequate samples.[5]

This method is an invasive method which also makes it inconvenient for the patients. Due to the various disadvantages that this method offers, it becomes a rather less preferred modality.

Salivary gland tumors are increasingly being evaluated prior to surgery using MRI since it can pinpoint their precise location, size, and surrounding tissues. [4], [6]

It is highly predictive of malignancy when certain MRI characteristics of parotid gland tumors obtained from conventional MRI, such as an ill-defined margin and low signal intensity (SI) on T2 weighted images (T2WIs), are present. [7]

To effectively supplement traditional MRI, apparent diffusion coefficient (ADC) values derived from diffusion-weighted imaging (DWI) data provide additional quantitative data regarding the random transport of water molecules in tissues. [8]

Based on this, it is safe to claim that MRI, together with its extra components, aids in the diagnosis of the tumor and pertinent traits needed to create a successful recovery plan.

This meta-analysis aims to ascertain the effectiveness of MRI versus the role of MRI with diffusion-weighted imaging.

## METHODOLOGY

### DATA COLLECTION

For the collection of the data, a search was done by two individuals using PubMed, Google Scholar, and Cochrane Library databases for all available relevant literature. We only considered Full - Text Articles written only in English, for this study.

The medical subject headings (MeSH) and keywords ‘Parotid carcinoma imaging’, ‘Result of Diffusion-weighted imaging (DWI) MRI Scan in parotid carcinoma diagnosis’, and ‘MRI in Parotid carcinoma’ were used. References, reviews, and meta-analyses were scanned for additional articles.

### INCLUSION AND EXCLUSION CRITERIA

Titles and abstracts were screened, and Duplicates and citations were removed. References of relevant articles were reviewed for possible additional literature. Papers with detailed patient information and statically proved and well-calculated results were selected.

We searched for papers that show more accurate diagnoses, where procedures considered were MRI with DWI and Only MRI in the diagnosis of parotid carcinoma.

The inclusion criteria were as follows:; (1) Literature that provided information about the accurate diagnosis with MRI(DWI) and only MRI ; (2) papers published in English; (3) papers with appropriate patient criteria.

The exclusion criteria were: (1) articles that were not full text, (2) unpublished articles, and (3) non-English articles.

### DATA EXTRACTION

Each qualifying Literature was independently assessed by two reviewers. Each article was examined for the number of patients, age, sex, procedure modality, comorbidity, and incidence of the predecided complications. Further debate or consideration with the author and a third party was used to resolve conflicts. This study’s quality was assessed using the modified Jadad score. In conclusion, As stated by PRISMA, a total of 8 RCTs with a total of 944 patients were selected, out of which 3 RCTs with a total of 356 patients were selected for MRI and 5 RCTs with a total of 588 patients were selected for MRI+DWI study.

### ASSESSMENT OF STUDY QUALITY

Two writers independently assessed the quality of each included RCT. This test consists of 10 questions, each with a score between 0 and 2, with 20 being the maximum possible overall score. Two authors evaluated each article independently based on the above criteria. The interobserver bias for study selection was determined using the weighted Cohen’s kappa (K) coefficient. For deciding the bias risk for RCTs, we also employed the Cochrane tool. No assumptions were made about any missing or unclear information. there was no financial aid involved in collecting or reviewing data.

### STATISTICAL ANALYSIS

The statistical software packages RevMan (Review Manager, version 5.3), SPSS (Statistical Package for the Social Sciences, version 20), Google Sheets, and Excel in Stata 14 were used to create the statistical analyses. The data was created and entered into analytic software [9]. Fixed- or random-effects models were used to determine Sensitivity, Specificity, positive predictive value (PPV), diagnostic odds ratios (DOR), and relative risk (RR) with 95 percent confidence intervals to examine critical clinical outcomes (CIs). Diagnosis accuracy and Younden index were calculated for each result. Individual study sensitivity and specificity were plotted on Forest plots and in the receiver operating characteristic (ROC) curve. The forest plot and Fagan’s Nomogram were used to explain the sensitivity and specificity of different papers.

### BIAS STUDY

The risk of bias was estimated by using the QUADAS-2 analysis tool. This tool includes 4 domains as Patient selection, Index test, Reference standard, Flow of the patients, and Timing of the Index tests.

## RESULT

### MRI

Here, Table 1 describes all the description of papers used for the study. As the result calculated above, in the forest chart (figure 2), the comparison of the sensitivity and specificity of different papers can be seen. The same can be seen depicted in the SROC curve(figure3). A total of 3 RCTs with 356 patients were selected for the study. The value of True positive is 170, True Negative is 112, False negative is 45, and False Positive is 29. With a confidence interval of 95%, Sensitivity, specificity, and Positive Predictive values were also calculated and summarised. A summary of this is available in Figure 2. The Sensitivity of the MRI is 0.790 with a CI of 95% in a range of (0.772 to 0.809) the mean being (0.0183). The Specificity of the MRI is 0.794 with a CI of 95% in a range of (0.613 to 0.975) the mean being (0.181). The positive predictive value (PPV) for the MRI is 0.854 with a CI of 95% in a range of (0.812 to 0.895) the mean being (0.0416).

**Table 1:** Table of Description of papers.

| Author Name and year | Duration | Mean Age | Sample size | Study Design | Index test | Reference test | Favorable towards |
| --- | --- | --- | --- | --- | --- | --- | --- |
| Can Wang 2021[5] | January 2004 – December 2017 | 57.6 | 113 | Retrospective review | CT + MRI | Surgery and Histopathologic examination | CT + MRI |
| Hao Hu 2021[10] | November 2019 – October 2020 | 50.4 | 58 | Retrospective review | 3D pseudo-continuous arterial spin labeling MRI | Surgery | MRI |
| Koshi Ikeda 1996[11] | January 1990 – August 1993 | - | 82 | Retrospective review | MRI | Histopathologic examination | MRI |
| Hidetake Yabuuchi 2008[12] | April 2004 – September 2007 | 61 | 47 | Retrospective review | MRI DWI + MRI DCE | Surgery + Histopathologic examination | MRI DWI + DCE |
| Irfan Celebi 2013[13] |  | 47 | 75 | Introspective | MRI DWI | Biopsy/FNAC | MRI DWI |
| J. Lechner Goyault 2010[14] | April 2005 – February 2008 | 59.4 | 60 | Retrospective review | MRI DWI | Histopathologic examination | MRI DWI |
| Xiaofeng Tao 2017[15] | October 2004 – December 2012 | 52 | 148 | Retrospective review | MRI + DWI + DCE | Surgery + Histopathologic examination | MRI + DWI + DCE |
| Ying Yuan 2016[16] | January 2010 – December 2014 | 48.4 | 207 | Retrospective review | MRI + DWI + DCE | Histopathologic examination | MRI + DWI |

**Figure 1.**
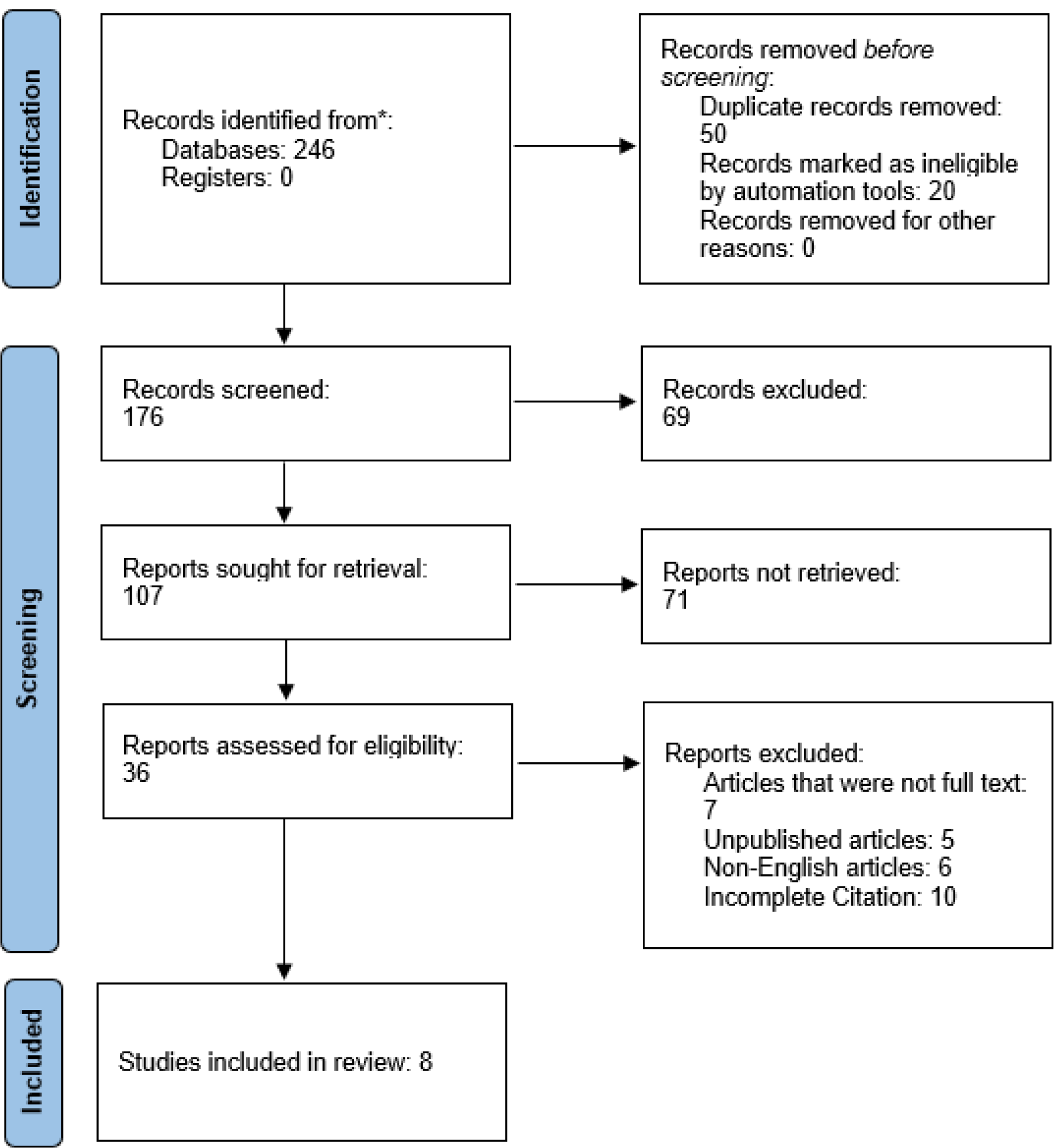
PRISMA Flowchart.

**Figure 2:**
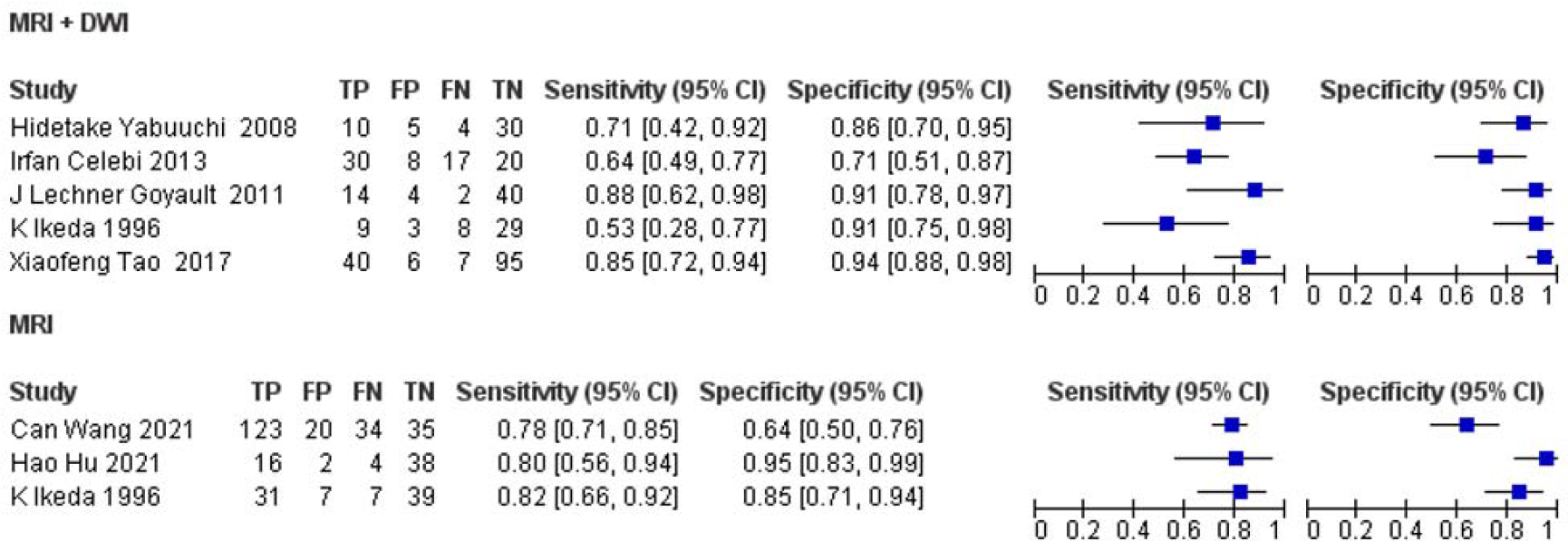
Forest chart for MRI+DWI and MRL

The summary of the ROC curve is illustrated in Figure 3. It describes that the area under the ROC (AUC) for MRI is 0.7925 and the overall diagnostic odds ratio (DOR) is 14.59. It also describes the Diagnostic Accuracy and The Younden index. which are 0.792 and 0.585 respectively.

**Figure 3:**
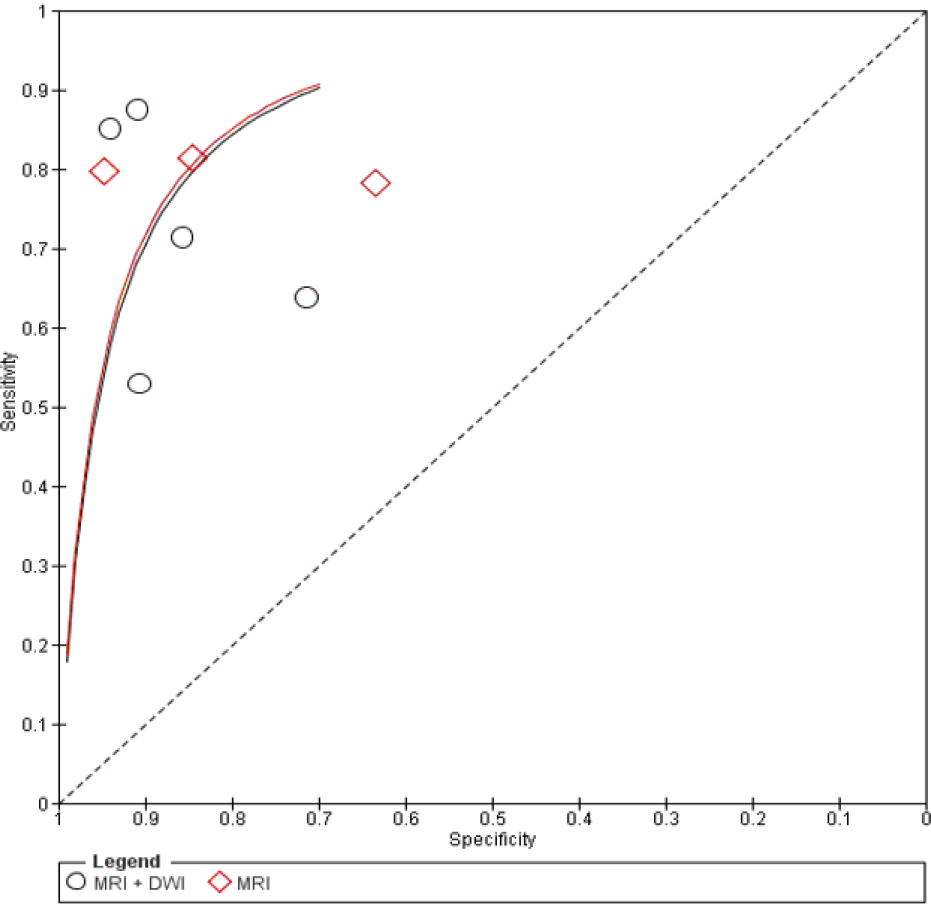
Summary of SROC plot

Figure 4 shows the summary of Fagan plots analysis for all the studies of MRI, it shows a Prior probability of 60% (1.5); a Positive likelihood ratio of 3.84; a probability of post-test 85% (5.9); a Negative likelihood ratio of 0.26, and a probability of post-test 28% (0.4).

**Figure 4:**
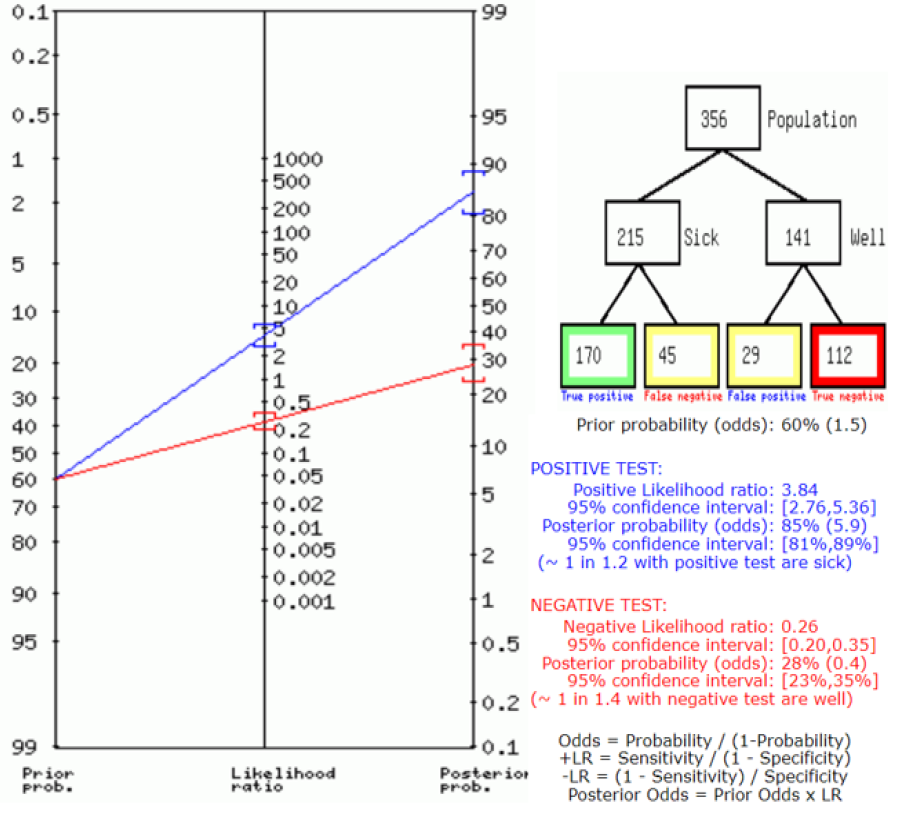
Fegan’s analysis for MRI

### MRI+DWI

Above, Table 2 Describes all the description of papers used for the MRI+DWI study. As the result showed above, in the forest chart (Figure 2), the comparison of the sensitivity and specificity of different papers can be seen. The same is illustrated in the SROC curve (figure 3). A total of 5 RCTs with 588 patients were selected for the study. The value of True positive is 215, True Negative is 214, False negative is 133, and False Positive is 26. With a confidence interval of 95%, Sensitivity, specificity, and Positive Predictive values were calculated. A summary of this is available in Figure 2. The Sensitivity of the MRI+DWI is 0.618 with a CI of 95% in a range of (0.498 to 0.737); the mean being (0.119). The Specificity of the MRI+DWI is 0.892 with a CI of 95% in a range of (0.813 to 0.97) the mean being (0.079). The positive predictive value (PPV) for MRI+DWI is 0.892 with a CI of 95% in a range of (0.801 to 0.984) the mean being (0.091).

The summary of the ROC curve is described in Figure 3. It shows that the area under the ROC (AUC) for the MRI+DWI is 0.75474 and the overall diagnostic odds ratio (DOR) is 13.305. Also, The Diagnostic Accuracy and Younden index are 0.73 and 0.509 respectively.

Figure 5 describes the summary of Fagan plots analysis for all the studies of MRI+DWI, it shows a Prior probability of 59% (1.5); a Positive likelihood ratio of 5.7; a probability of post-test 89% (8.3); a Negative likelihood ratio of 0.43, and a probability of post-test 38% (0.6).

**Figure 5:**
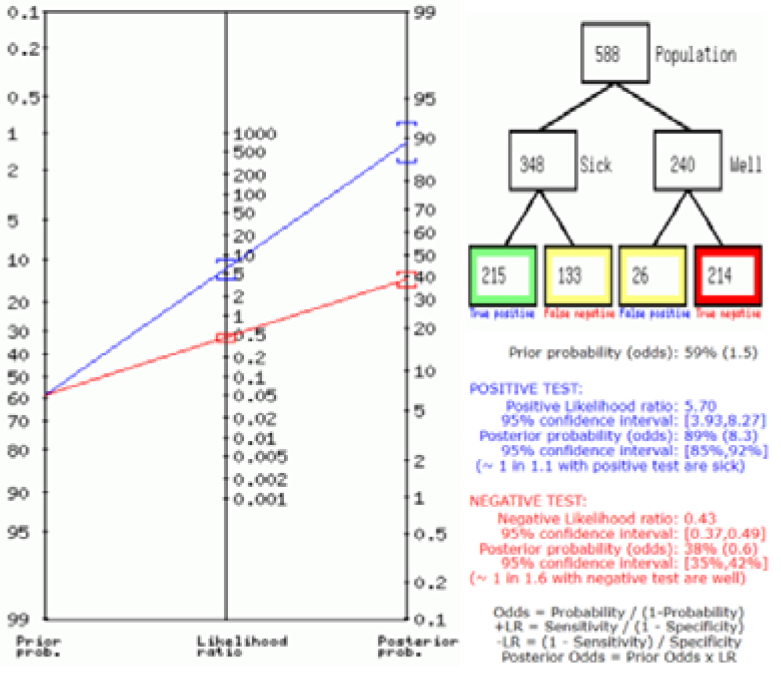
Fegan’s analysis for MRI-DWI

### Bias Study

#### Publication Bias

The summary of publication bias for MRI+DWI and MRI is shown in (Figure 6 and Figure 7). For the publication bias, In, patient selection, bias was low in 6 studies and unclear in 3. In the index test, it was low in 9 studies. While the reference standard was low in 4, unclear in 3, and high in 2. The flow and timing were low in 9. The applicability concerns in patient selection were low in 7 and high in 2. The index test was low in 7 and unclear in 2. The reference standard was low in 2, unclear in 2, and high in 5.

**Figure 6:**
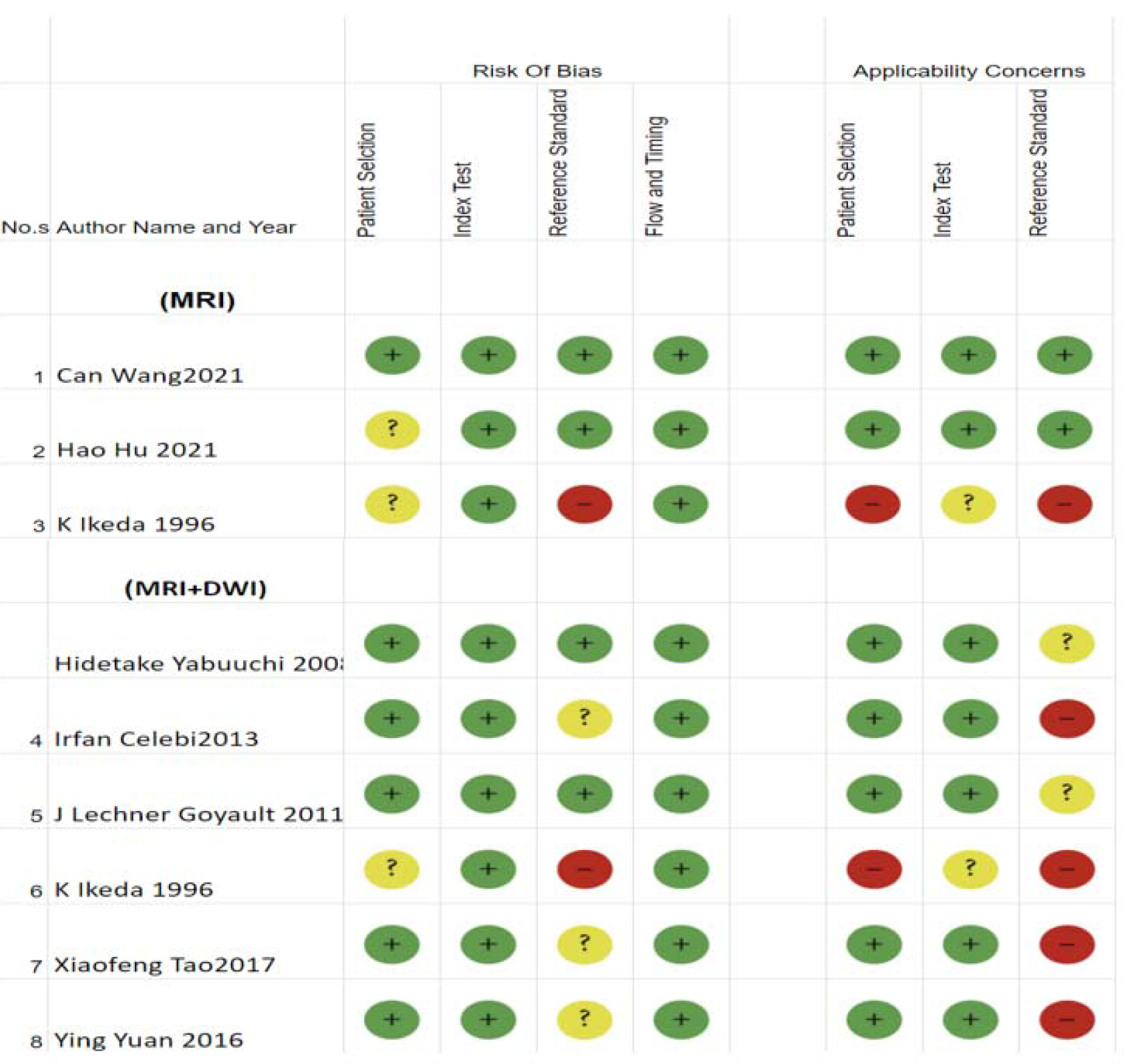
Table of Bias study

**Figure 7:**
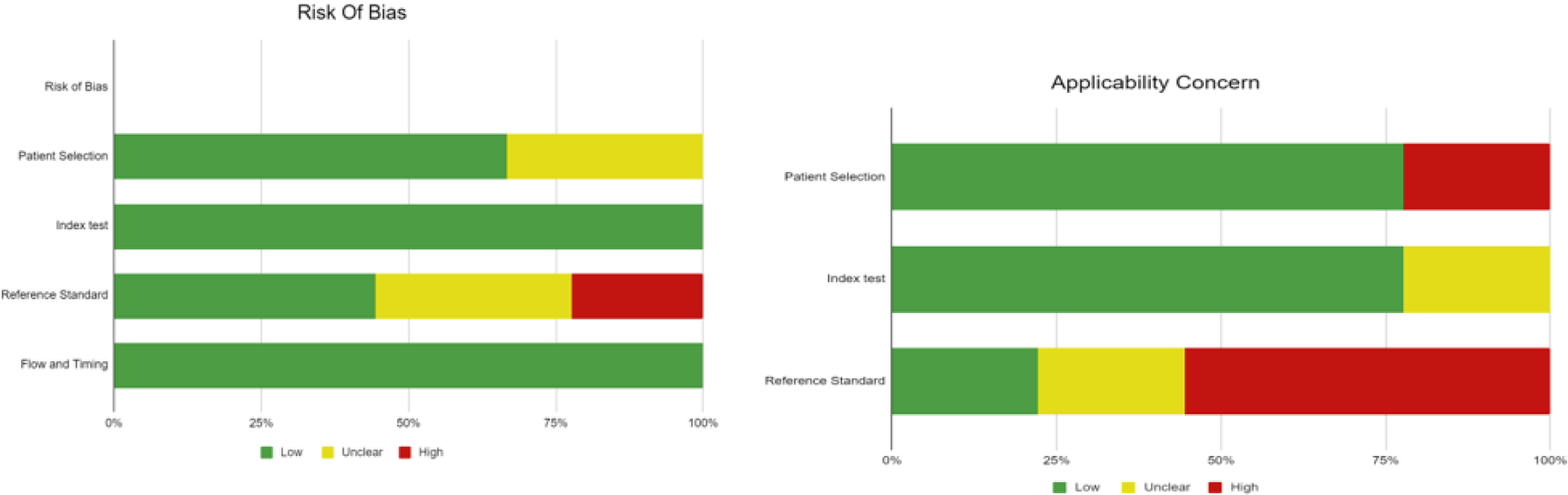
Summary of Bias study

## Discussion

As different surgical techniques are used for benign and malignant lesions, the Preoperative determination of malignancy in the parotid gland tumors is clinically important [17]. Some MRI features obtained from conventional MRI for parotid gland lesions, such as ill-defined margin and low signal intensity on T2WI, are major indicators of malignancy [7], [18]. However, some researchers hold the opposite conclusion and the sensitivities and specificities of these results have been identified as low and significantly overlapping[19], [20]. DWI has been verified to be a rather beneficial technique for assessing the characteristics of head and neck tumors.[21]–[25].

For Parotid gland neoplasms, Functional MRI (DWI) can help to identify an initial diagnosis of a benign tumor rather than a malignant one, especially in those cases where the tumor is well-defined. Pleomorphic adenomas classically involve a significant amount of myxomatous tissue, which is the root for the less restriction of water diffusion, high ADCs, and high signal intensity on the ADC map when compared to metastatic nodes or primary salivary gland carcinomas. Warthin tumors, in contrast, may include ample amounts of densely packed lymphoid cells, which accounts for their lower ADCs, which overlap with those of malignancy; however, Warthin tumors are usually highly vascular compared with pleomorphic adenomas and malignant salivary gland tumors.[26] Consequently, IVIM can help to differentiate benign from malignant tumors by using high Diffusion to reflect higher pseudo-diffusion in the microcirculation in Warthin tumors and high Diffusion to reflect less restricted pure diffusion in pleomorphic adenoma.

Therefore, we tried to compare the diagnostic accuracy of the simple MRI with DWI MRI for parotid gland neoplasms. The Sensitivity of the MRI is 0.790 (CI95%,0.772-0.809) and the Specificity of the MRI is 0.794 (CI95%,0.613-0.975). and the Sensitivity of the MRI+DWI is 0.618 (95%,0.498-0.737) and the Specificity of the MRI+DWI is 0.892 (95%,0.813-0.97). There is very little improvement in Specificity here but concerns about resource usefulness and technicality regarding the procedure remain. Here we also describe the diagnostic odds ratio and Youden index for both, it shows very little or no improvement.

Therefore it is safe to say that neither simple MRI nor DWI MRI is independently and thoroughly helpful for accurate diagnosis and early detection of Parotid malignancy or in distinguishing benign lesions from malignant ones. However, using DWI MRI and Simple MRI together for the detection and differentiation of benign and malignant lesions would be very accommodating. We recommend for more powerful modalities like DCE-MRI should be incorporated for the detection and defining of the margins of parotid gland lesions before surgery. A combination of DCE-MRI with DWI has been confirmed to advance the MRI performance for characterizing benign and malignant parotid gland tumors and the difference in distinguishing histological types of benign tumors.[14] [12].

## Limitations

This meta-analysis did not add the other articles from different languages other than English. The ADC value varies depending on equipment and hospital imaging method which is why the results regarding ADC and TIC are not liberally accurate. We were not able to quantify the patient age factor as it changes the parotid gland signals in MRI, so this suggests dividing future studies into the different age groups for better results and quantification process.

## Conclusion

In conclusion, if parotid gland tumors are shown on MRI as being of irregular shape malignancy should be strongly considered. In addition to simple MRI or Single DWI MRI, A combination of both would detect higher lesions and it should be helpful for staging and early diagnosis purposes only. For perfect anatomical understanding and to plan surgical approaches, more studies, and research should be done exploring newer and more powerful modalities like DCE MRI.

## Data Availability

All data produced in the present work are contained in the manuscript

